# Circulating pancreatic enzyme levels are a causal biomarker of type 1 diabetes

**DOI:** 10.1101/2024.08.08.24311619

**Authors:** Ruth M Elgamal, Rebecca L. Melton, Joshua Chiou, Carolyn W McGrail, Kyle J Gaulton

**Affiliations:** Biomedical Sciences Graduate Program, UC San Diego, La Jolla CA; Department of Pediatrics, UC San Diego, La Jolla CA; Pfizer Research and Discovery, Pfizer Inc., Cambridge, MA

**Author notes:** Correspondence to Kyle J. Gaulton.

## Abstract

Novel biomarkers of type 1 diabetes (T1D) are needed for earlier detection of disease and identifying therapeutic targets. We identified biomarkers of T1D by combining plasma *cis* and *trans* protein QTLs (pQTLs) for 2,922 proteins in the UK Biobank with a T1D genome-wide association study (GWAS) in 157k samples. T1D risk variants at over 20% of known loci colocalized with *cis* or *trans* pQTLs, and distinct sets of T1D loci colocalized with immune, pancreatic secretion, or gut-related proteins. We identified 23 proteins with evidence for a causal role in using pQTLs as genetic instruments in Mendelian Randomization which included multiple sensitivity analyses. Proteins increasing T1D risk were involved in immune processes (e.g. *HLA-DRA*) and, more surprisingly, T1D protective proteins were enriched in pancreatic secretions (e.g. *CPA1*), cholesterol metabolism (e.g. *APOA1*), and gut homeostasis. Genetic variants associated with plasma levels of T1D-protective pancreatic enzymes such as CPA1 were enriched in *cis*-regulatory elements in pancreatic exocrine and gut enteroendocrine cells, and the protective effects of CPA1 and other enzymes on T1D were consistent when using instruments specific to acinar cells. Finally, pancreatic enzymes had decreased acinar expression in T1D, including CPA1 which was altered prior to onset. Together, these results reveal causal biomarkers and highlight processes in the exocrine pancreas, immune system, and gut that modulate T1D risk.

## Introduction

Type 1 diabetes (T1D) is a complex disorder characterized by autoimmune destruction of insulin-producing beta cells in the pancreatic islets and subsequent hyperglycemia. Identifying biomarkers for T1D can inform disease prediction and diagnosis, selection of individuals for therapies, and identification of new therapeutic targets. Seroconversion to islet-specific autoantibodies precedes dysglycemia and the development of T1D^1^ in the majority of cases, and therefore represents a robust biomarker of T1D. It is unknown; however, whether islet-specific autoantibodies play a role in disease pathogenesis directly and thus may not represent modifiable factors to prevent T1D^2^. Furthermore, recent work has argued that beta cell dysfunction already exists at the time of seroconversion^3,4,5,6^. There is therefore a need for novel biomarkers of T1D for both earlier detection of disease as well as to identify modifiable factors that prevent disease.

Identifying robust biomarkers of T1D, however, has thus far had limited success. Cellular biomarkers such as infiltrating antigen-specific and cytotoxic T-cells are tissue-localized to the site of autoimmune attack in the pancreas and are difficult to distinguish in circulating blood^7,8^. Other studies have identified altered circulating plasma levels of proteins and metabolites preceding T1D onset, although in limited sample sizes^9^. Furthermore, few studies have assessed whether circulating biomarkers are causally linked to the development of T1D^10^. Mendelian randomization (MR) is a technique that can assess whether exposures such as circulating protein levels have a causal effect on T1D using human genetic association data^11^. Studies using MR have revealed exposures with both evidence for causality in T1D such as childhood BMI^12,13^ and levels of several circulating proteins^14^ and clarified those unlikely to play a causal role in T1D such as vitamin D levels^15^ and pancreatic volume^16^. Statistical colocalization is an orthogonal technique that can identify exposures associated with T1D risk at individual loci based on shared causal variants. This method has been applied to T1D primarily in the context of gene expression QTLs. For both approaches a key limitation to date has been the availability of studies of sufficient scope and sample size which are needed to establish robust relationships to disease.

In this study, we evaluated circulating proteins for a causal role in T1D risk by utilizing QTLs for plasma levels of 2,922 proteins from 35k individuals in the UK Biobank Pharma Proteomics Project (UKB-PPP)^17^. We performed colocalization and MR analyses of plasma protein QTLs (pQTLs) with a T1D genome-wide association study (GWAS) in 157k samples^1^. We then characterized potential mechanisms of proteins with evidence for a causal role in T1D using genetic association and single cell genomics data.

## Results

### Colocalization of T1D loci with plasma protein QTLs

We identified T1D risk loci genome-wide with evidence for affecting circulating protein levels. We utilized genome-wide association data for T1D in a meta-analysis of 157k samples (**Supplementary Table 1**), as well as *cis* and *trans* quantitative trait loci for plasma levels of 2,922 unique proteins (pQTLs) in 35k samples from the UKB-PPP project and performed statistical colocalization. In total, across 92 known T1D loci, 20 (21.7%) were colocalized with plasma pQTLs (PPH4 >0.8, r^2^ between lead variants >0.5) using coloc^18^ (**Fig. 1a, Supplementary Table 2**). Most loci were colocalized with pQTLs for a single protein, although three loci (*SH2B3, FUT2, IRF1*) colocalized with more than 50 proteins each (**Fig. 1a**). The majority of colocalized pQTLs were for *trans* signals.

**Fig. 1:**
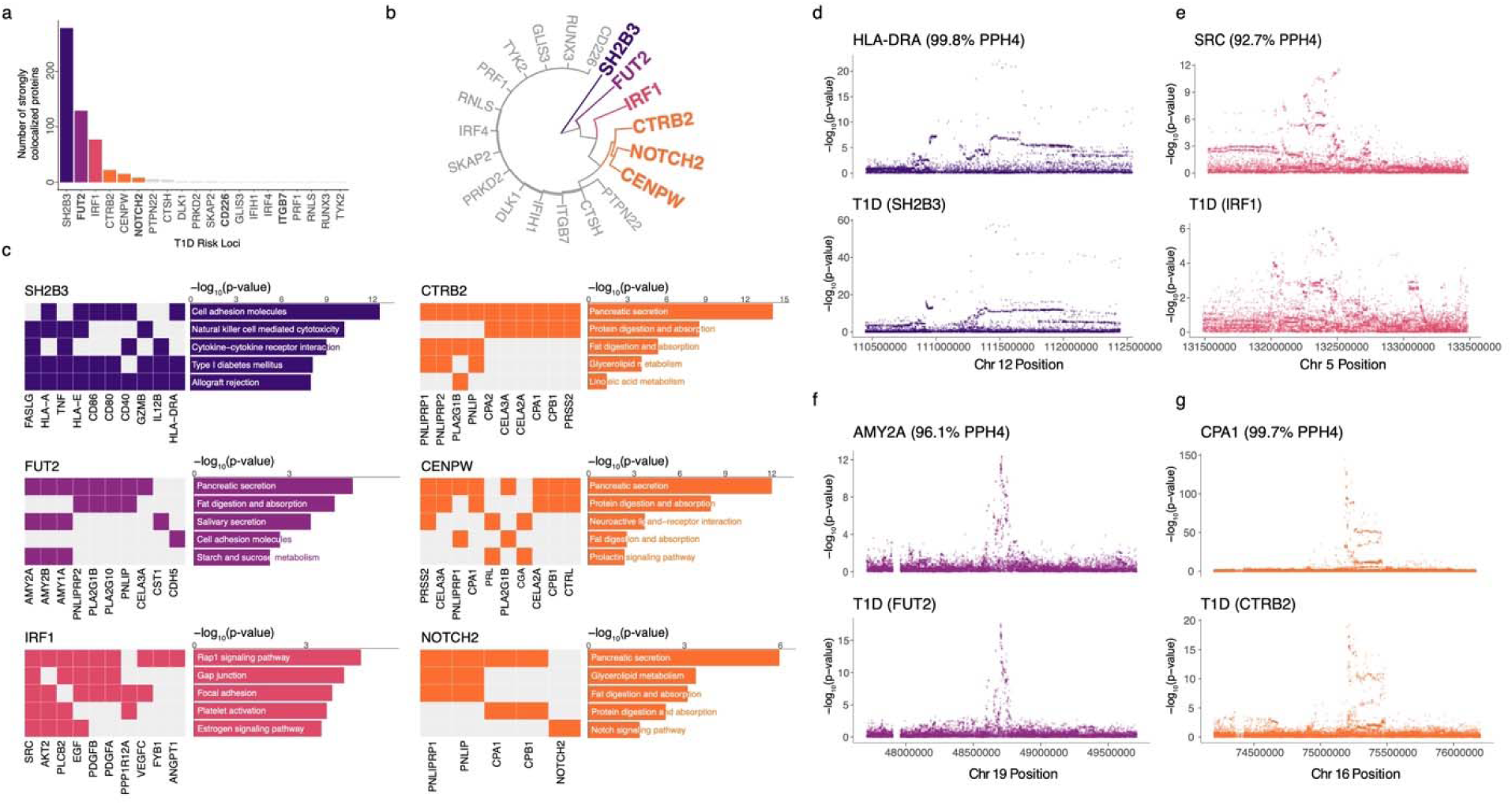
Colocalization of UKB-PPP proteins with T1D reveals mechanisms at T1D risk loci. **a)** Number of strongly colocalized UKB-PPP proteins (PPH4 > 80%) at each T1D risk locus that had at least one colocalized protein. Loci are colored by their hierarchical clustering. Bolded loci strongly colocalized to at least one *cis* pQTL. **b)** Circular dendrogram of hierarchical clustering of T1D risk loci based on their colocalization across UKB-PPP proteins. **c)** KEGG pathway enrichment of strongly colocalized proteins at T1D risk loci including pathway membership of top 10 proteins per locus. **d-g)** Colocalization locus plots of **d)** HLA-DRA plasma pQTL at the SH2B3 T1D risk locus, **e)** SRC plasma pQTL at the IRF1 T1D risk locus **f)** AMY2A plasma pQTL at the FUT2 T1D risk locus and, **g)** CPA1 plasma pQTL at the CTRB2 T1D risk locus.

Colocalizations with plasma pQTLs provided novel, and in some cases unexpected, insight into the potential mechanisms of risk variant activity at many loci. For example, the *RNLS* locus is colocalized with plasma levels of colipase (CLPS), which is a co-factor of pancreatic lipase produced specifically in pancreatic acinar cells. Risk variants at *RNLS* are also colocalized with an expression QTL for the *PTEN* gene in pancreas in the GTEx database^19^, supporting a potential function of this locus in pancreatic acinar cells. The *GLIS3* locus is colocalized with plasma levels of thyroglobulin (TG), which is produced in follicular cells of the thyroid, and *GLIS3* is an essential transcriptional regulator of thyroid hormone production^20,21^. Although the *RNLS* and *GLIS3* loci are both implicated in beta cell function from previous studies^22,23^, pQTL colocalization results suggest risk variants at these loci may also affect other systems.

We next performed hierarchical clustering of all T1D risk loci with at least one colocalized pQTL to determine whether loci could be grouped based on their colocalization profiles (**Fig. 1b**). The three loci with many colocalizations *SH2B3*, *FUT2*, and *IRF1* had largely non-overlapping sets of colocalized proteins. SH2B3, in particular, had over 200 strongly colocalized proteins with many immune-related pQTLs sharing the same direction of effect as T1D risk. Three other loci *CTRB2*, *CENPW*, and *NOTCH2* clustered together, where four proteins strongly colocalized with all three loci and an additional seven proteins colocalized with two of the three loci. Proteins colocalizing with these three loci included multiple pancreatic enzymes (CPA1, CPB1, PNLIP, CELA2A/3A) as well as pancreatic enzyme-associated proteins (PNLIPRP1, a pancreatic lipase related protein, and SERPINI2, a serpin peptidase inhibitor). Interestingly, T1D protective alleles at these loci were correlated with increased pancreatic enzyme levels. The *FUT2* locus was also colocalized with multiple pancreatic enzymes, including those not colocalized with other T1D loci such as amylase (AMY2A/B).

Pathway enrichment of shared proteins for each T1D risk locus with more than two colocalized pQTLs (**see Methods**) revealed further insight into potential mechanisms of these loci (**Fig. 1c**). For example, pQTLs colocalized with the *SH2B3* locus were enriched for cytokine-cytokine receptor interactions (FDR=5.65×10^−8^) and viral interactions with cytokine receptors (FDR=2.06×10^−7^), *SH2B3* and *FUT2* were enriched for cell adhesion molecules (FDR=5.10×10^-11^, FDR=1.93×10^−3^), and *IRF1* was enriched for RAP1 and RAS signaling (FDR=6.43×10^−3^, FDR=1.53×10^−3^). By comparison, colocalized pQTLs at the *CTRB2, CENPW, NOTCH2* loci, as well as *FUT2*, were all enriched for pancreatic enzyme-related pathways such as pancreatic secretions (FDR=2.26×10^−13^, FDR=2.37×10^−11^, FDR=1.45×10^−5^, FDR=9.90×10^−6^) and protein digestion and absorption (FDR=5.07×10^−8^, FDR=1.22×10^−7^, FDR=1.31×10^−3^).

Together these results reveal a large proportion of T1D loci colocalized with plasma *cis* and *trans* pQTLs and implicate cytokine receptor interactions and pancreatic enzymes, among other processes such as gut homeostasis, in T1D risk at individual loci.

### Plasma proteins with a causal role in T1D using Mendelian randomization

We next assessed whether circulating levels of specific proteins may be causally linked to T1D pathogenesis. For each protein, we defined genetic instrumental variables consisting of all independent *cis* or *trans* QTL signals strongly associated with plasma levels of the protein (**see Methods**). We then tested instruments for association to T1D using the inverse variance weighted (IVW) method for Mendelian randomization (MR)^24^.

In total, we identified 23 proteins with evidence for a causal role in T1D when using combined *cis* and *trans* pQTLs (FDR<0.1) **(Fig. 2a, Supplementary Table 3)**. Among these 23 proteins, increased circulating levels of 12 proteins were protective for T1D, whereas increased circulating levels of 11 proteins increased risk of T1D. Proteins with the largest protective effects on T1D included DXO, PRSS2, TGFA, and CPA1, while proteins with the largest risk effects included HLA-DRA, CASP8, and APBB1IP. The 12 protective proteins were significantly enriched for pathways related to cholesterol metabolism (LRPAP1, APOC1, APOA1; FDR=3.49×10^−3^), pancreatic secretions (PNLIPRP1, CPA1, PRSS2; FDR=5.75×10^−3^), fat digestion/absorption (PNLIPRP1, APOA1; FDR=2.34×10^−2^), and protein digestion/absorption (CPA1, PRSS2; FDR=8.32×10^−2^) (**Fig. 2b**). Several other proteins showing protective effects on T1D, ALPI and TGFA, are involved in gut homeostasis. By comparison, the 11 proteins that increased T1D risk were enriched in pathways for antigen processing and presentation (HLA-DRA, LGMN; FDR=1.14×10^−1^) and other immune-related processes (**Fig. 2c**). Another risk-increasing protein APBB1IP is involved in T cell activation through contact with antigen presenting cells.

**Fig. 2:**
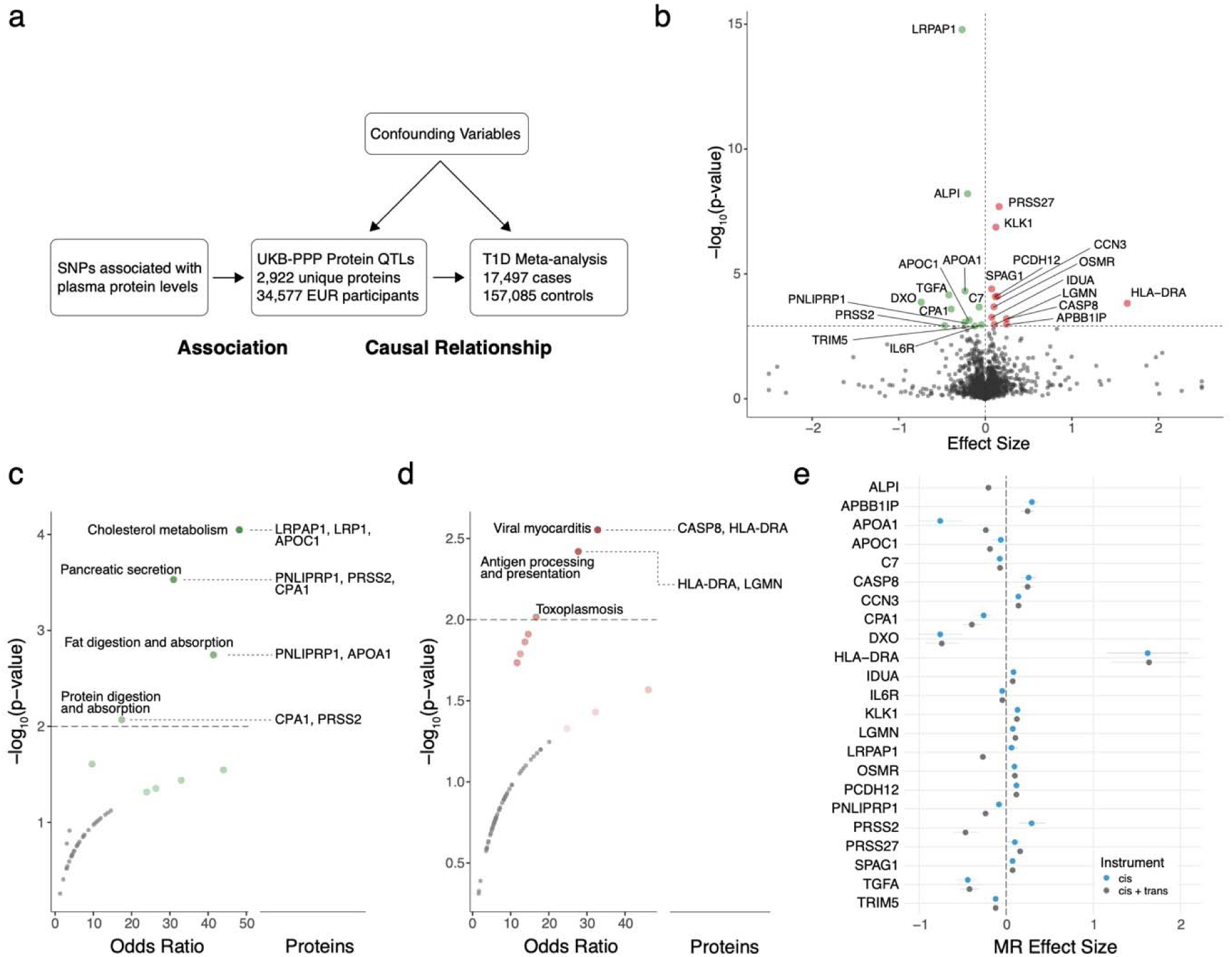
Mendelian randomization of UKB-PPP proteins with T1D identifies causal proteins. **a)** Study design for Mendelian randomization of UKB-PPP proteins with T1D outcome to determine causal proteins in T1D. **b)** Volcano plot of Mendelian randomization effect sizes and - log_10_(p-values) across 2,922 UKB-PPP proteins. Proteins in green have a protective causal effect (beta < 0) while proteins in red have a risk-inducing causal effect (beta > 0). Effect sizes for non-significant proteins were capped at -2.5 and 2.5. **c)** Odds ratios and -log_10_(p-values) of KEGG pathway enrichment of protective causal proteins. **d)** Odds ratios and p-values of KEGG pathway enrichment of risk-inducing causal proteins. **e)** Forest plot of Mendelian randomization effect sizes for *cis* and *trans* QTL instruments compared to *cis* QTL only instruments. Error bars represent standard error of the mean for Mendelian randomization effect sizes.

We next performed sensitivity analyses of the 23 proteins with evidence for a causal role in T1D. To determine whether significant effects could potentially be explained by horizontal pleiotropy, we calculated the MR Egger intercept for each set of instruments. Overall, no instrument showed significant, or even nominally significant, evidence for pleiotropy **(Supplementary Table 4)**. Next, as the inclusion of outliers in MR instruments can indicate pleiotropic bias, we used MR-PRESSO to detect outlying variants in pQTL instruments as well as re-evaluate MR after outlier removal^25^. We identified outlying variants in the instruments of 9 proteins and after re-analysis with outlier correction, 5 of the 9 (CPA1, DXO, KLK1, OSMR, and PRSS2) still had significant effects on T1D **(Supplementary Table 5, Supplementary Fig. 1a)**. Finally, we performed leave-one-out analyses to determine the impact of individual variants in each instrument on MR. All but one protein (ALPI) had consistent effect sizes when iteratively excluding individual variants, indicating robust instruments **(Supplementary Fig. 1b, Supplementary Table 6)**. For ALPI, leave-one-out analysis indicated that the causal effect was driven by a single variant at the *FUT2* locus. In total, these results reveal proteins with consistent effects for causal effects on T1D after multiple sensitivity analyses.

The inclusion of *trans* signals in pQTL instruments may result in variants with indirect effects on a protein. Therefore, we evaluated whether proteins had consistent effects on T1D using instruments consisting of only *cis* pQTLs. Of the 23 proteins with significant effects in MR using combined *cis* and *trans* pQTLs, all but one (ALPI) had had at least one independent *cis* QTL.

When using instruments containing only *cis* pQTLs in MR, almost all proteins (87%, 20/22) showed directionally consistent effects on T1D, of which 17 still remained significant at FDR<0.10 **(Fig. 2d, Supplementary Table 7)**. The small number of proteins where *cis* pQTL instruments did not have consistent effects on T1D included PRSS2, which had the opposite effect on T1D compared to the combined *cis* and *trans* pQTLs, and LRPAP1, which had little to no effect on T1D **(Fig. 2d, Supplemental Fig. 1c).**

In total, across multiple sensitivity analyses and genetic instruments, 14 proteins showed consistent causal effects on T1D. These included pancreatic proteases (CPA1, KLK1), other enzymes (CASP8, IDUA), the apolipoprotein APOA1, cytokine receptors IL6R, OSMR, complement factor C7, as well as proteins involved in T cell-APC interactions (APBB1IP, HLA-DRA) and cell adhesion (CCN3, PCDH12). Overall, circulating proteins causally associated with T1D implicate diverse processes including pancreatic secretions, digestion, antigen presentation, and immune activity in T1D.

### Enrichment of plasma protein QTLs for cell type-specific regulatory elements

Levels of a protein in plasma are likely are result of multiple intra- and extra-cellular processes including production, secretion, and degradation, and understanding the processes underlying plasma levels may provide insight into how they contribute causally to T1D. As most genetic variants associated with protein levels are in non-coding regions and likely affect gene regulation, intersection of associated variants with cell type-specific epigenomic maps can help reveal cell types driving plasma levels of a protein.

We tested for genome-wide enrichment of plasma pQTLs for T1D-causal proteins in *cis*- regulatory elements (cREs) for 110 adult cell types using stratified LD score regression^26–28^. In total, there were 188 significant (FDR<0.10) links between pQTLs and cell types **(Supplementary Table 8)**. Enriched cell types provided insight into processes underlying plasma levels of many proteins. For example, pancreatic enzymes CPA1, PNLIPRP1, and PRSS2 were significantly enriched in cREs in pancreatic acinar and ductal cells, where they are produced and secreted into the gut, as well as gut enterocyte and colon epithelial cells (**Fig. 3a- c, Supplementary Fig. 2a**). Interestingly, the enzyme KLK1, although highly expressed in the pancreas, had distinct enrichment patterns including lung (e.g. exocrine club cells), epidermal, and esophageal endothelium cREs. In other examples, HLA-DRA was significantly enriched for cREs in macrophages, where it is expressed, as well as, interestingly, acinar cells. Complement protein C7 was enriched in cREs in hepatocytes and macrophages, where complement proteins are produced^29,30^, as well as endothelial cells where it mediates inflammation^31^. Finally, TGFA, a growth factor that induces angiogenesis, was enriched in pericytes, the supporting cells around capillaries^32,33^, endothelial cells, and mast cells.

**Fig. 3:**
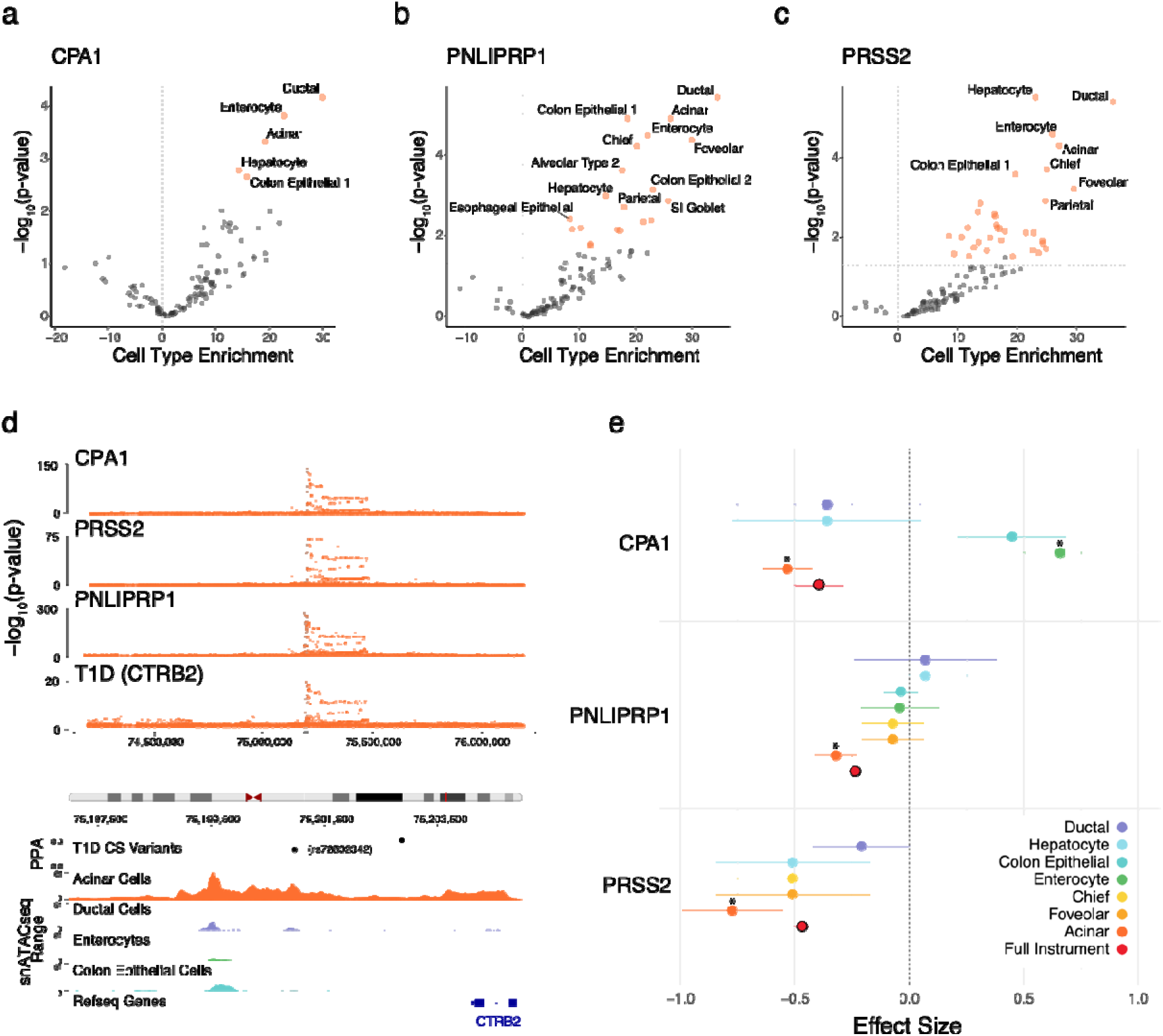
Pancreatic enzyme plasma protein QTLs are enriched in acinar-specific regulatory elements. Volcano plots of LD score regression cell type enrichments and corresponding -log_10_(p-values) for **a)** CPA1, **b)** PNLIPRP1, and **c)** PRSS2 pQTLs. Cell types with significant enrichments are annotated with green data points (FDR < 0.1). Dashed horizontal line represents a nominal p-value threshold of 0.05. **d)** Top– Colocalization of CPA1, PRSS2, and PNLIPRP1 pQTLs with the T1D risk locus at CTRB2. Bottom– Signal tracks representing chromatin accessibility at the CTRB2 locus in acinar, ductal, and colon epithelial cells from snATAC-seq in CATlas overlaid with fine-mapped T1D credible set variants. Risk variant rs72802342 lies within an acinar-specific peak. **e)** Mendelian randomization effect sizes of cell type-specific instruments generated from overlapping independent variants from CPA1, PNLIPRP1, and PRSS2 pQTLs with cell type cREs. Error bars represent standard error of the mean for Mendelian randomization effect sizes. Significant causal relationships are annotated with an asterisk (* FDR < 0.1).

Next, we annotated candidate causal variants at T1D risk loci colocalized with these proteins by leveraging cell type-specific cREs enriched for plasma pQTLs. The *CTRB1/2* locus is colocalized with pQTLs for multiple pancreatic enzymes (PRSS2, CPA1/2, CPB1, PNLIPRP1/2, CELA2A/3A), and genes at this locus encode for the digestive enzyme chymotrypsinogen. A candidate causal variant at *CTRB1/2* rs72802342 (T1D posterior probability=0.26) as well as several other lower probability variants (rs72802352, rs2039014912) overlapped cREs in acinar cells, and no other enriched cell types, supporting that altered acinar regulation underlies shared associations between T1D and pancreatic enzyme levels at this locus **(Fig. 3d, Supplementary Table 9)**. At *SH2B3*, which colocalized with serum levels of the MHC class II gene HLA-DRA, multiple candidate variants overlapped cREs active in macrophages and other immune cell types. Finally, at the *CENPW* locus, which colocalized with pancreatic enzymes (CPA1, PNLIPRP1, PRSS2), candidate variants overlapped cREs active in colon epithelial cells, gut enterocytes, and hepatocytes, suggesting cell types that could mediate risk at this locus.

We next determined whether the causal effects of pQTLs on T1D identified using MR were driven through specific cell types. For each of the 23 causal proteins, we generated ‘cell type-specific’ MR instruments by restricting the full instrument to just pQTL variants overlapping cREs in each cell type that was broadly enriched for pQTLs. For the pancreatic enzymes CPA1, PNLIPRP1, and PRSS2, pQTL variants overlapping pancreatic acinar cell cREs showed a significant protective effect on T1D (FDR=8.68×10^−6^, FDR=1.62×10^−2^, FDR=7.34×10^−3^), and in each case this protective effect was stronger compared to the full instrument **(Fig. 3e, Supplementary Table 10)**. Interestingly, CPA1 variants in gut enterocyte cREs also showed a significant causal relationship with T1D but in the opposite direction, suggesting that variants affecting CPA1 levels in different cell types may play distinct roles in disease.

We further used gene expression QTLs (eQTLs) to validate the tissue-specific effects of variants on plasma protein levels. We performed colocalization of *cis* pQTLs with single-tissue *cis* eQTLs from the GTEx project^19^ for the 23 causal proteins to identify individual tissues contributing to plasma *cis* pQTL signals. Overall, 14 out of 23 proteins strongly colocalized (PPH4>80%) with an eQTL for the gene encoding the respective protein in at least one GTEx tissue **(Supplementary Fig. 3a, Supplementary Table 11)**. Among pancreatic enzymes and enzyme-related proteins, PRSS2 was strongly colocalized to a pancreas *cis* eQTL, and pancreas was the tissue with the strongest evidence for colocalization for CPA1 and second strongest colocalization for PNLIPRP1 after sigmoid colon. Effect sizes in MR using pancreas *cis* eQTLs in GTEx for the pancreatic enzymes CPA1, PNLIPRP1, and PRSS2 were highly concordant (r^2^=0.99) with plasma *cis* pQTLs, where CPA1 had the largest causal effect in both *cis* pQTL and eQTL data **(Supplementary Fig. 3b-c, Supplementary Table 12)**.

Overall, these results highlight cell types that mediate plasma levels of many proteins with a causal link to T1D and, specifically, reveal acinar cells as a key driver of the protective effects of pancreatic enzyme levels on T1D.

### Changes in pancreatic enzyme activity in the pancreas in T1D

Multiple enzymes or enzyme-related proteins with causal links to T1D are primarily expressed and produced in pancreatic acinar cells, and therefore we finally determined whether genes encoding for these enzymes had altered expression in the pancreas in T1D progression. We therefore leveraged cell type-specific gene expression profiles from two previously generated single cell maps of scRNA-seq in pancreatic islets using data from the HPAP consortium^34^ and snRNA-seq in whole pancreas from the nPOD biorepository^35^. In total, across both maps, we utilized data from 82 donors which spanned multiple different stages of T1D progression including non-diabetic (n=40), single AAB+ (n=10), multiple AAB+ (n=10), recent-onset T1D (n=13) and long-duration T1D (n=9).

We determined changes in acinar expression in each gene across stages of T1D progression using a linear model **(see Methods**). CPA1, PNLIPRP1, and PRSS2 all showed decreased expression across stages of T1D progression, which is consistent with higher serum levels of enzymes being protective against disease, although only CPA1 had a significant effect (logFC per disease state=-0.220, FDR=3.23×10^−3^) **(Fig. 4a, Supplementary Table 13)**. We next determined changes in gene expression in each specific disease stage compared to non-diabetics. There was significantly (FDR=8.95×10^−2^) altered expression of CPA1 in single AAB+, as well as recent-onset and long-duration T1D, donors while PRSS2 had decreased expression in long-duration T1D only **(Fig. 4a-c, Supplementary Table 14)**. Notably, these two genes with significant decreases in expression both have protease activity.

**Fig. 4:**
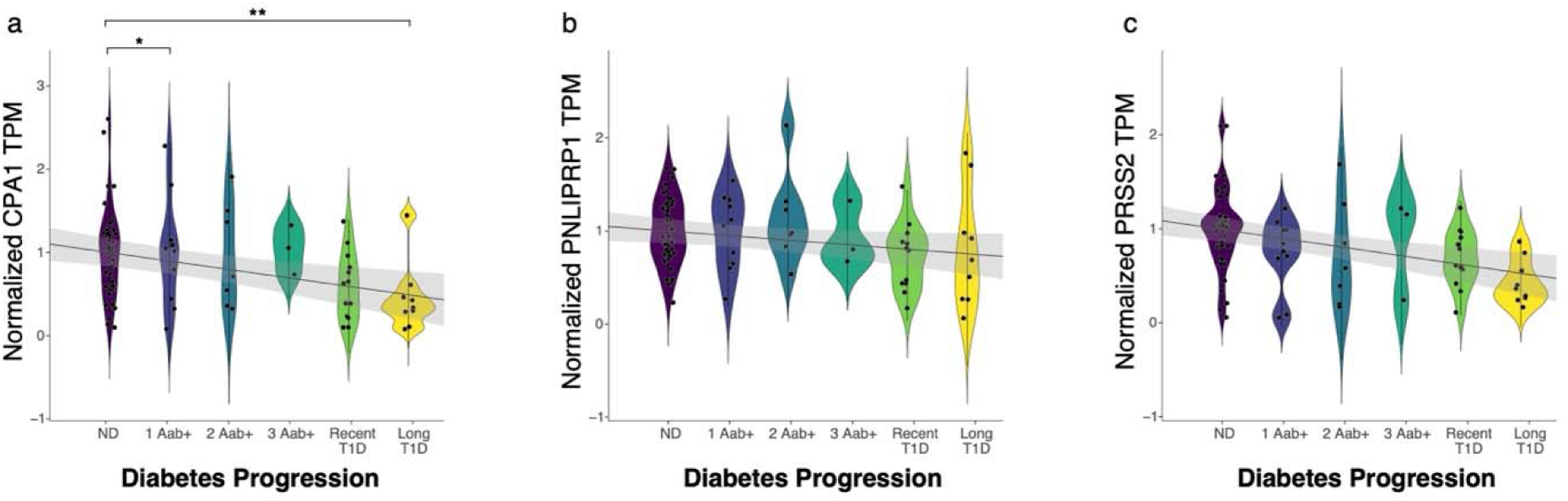
Acinar *CPA1* expression is decreased with T1D progression. **a-c)** Normalized TPMs as a function of T1D disease progression represented as a continuous variable from ND to long duration T1D for **a)** CPA1, **b)** PNLIPRP1, and **c)** PRSS2. Linear model and confidence interval are depicted for the relationship between pancreatic enzyme levels and disease progression. Error bars in violin plots represent standard deviation between donors. Pairwise comparisons between ND and diabetes stages were performed using DESeq2 and asterisks represent significant differences (FDR < 0.1; * < 0.1, ** < 0.01).

These results reveal that digestive proteases with a causal link to T1D have decreased expression in pancreatic acinar cells in T1D progression.

## Discussion

This study revealed that increased levels of several pancreatic enzymes due to altered regulation in the pancreas play a protective role in T1D development. In particular, CPA1 had robust support for a protective role in T1D across multiple different analyses and had altered expression in T1D progression, including in the early stages of disease, supporting that CPA1 is a novel biomarker of T1D. CPA1 encodes carboxypeptidase A1, a metalloprotease that cleaves dietary proteins and requires zinc for its enzymatic activity^36^. Although it is unknown how CPA1 activity protects against T1D, it is notable that pancreatic beta cells have high zinc levels which are required for the storage and secretion of insulin^37–39^. In addition, CPA1 is involved in zymogen inhibition, and therefore may protect against improper activation of enzymes in the pancreas which can lead to increased inflammation^36^. Supporting this, variants affecting CPA1 function are also linked to early-onset chronic pancreatitis and pancreatic cancer^40,41^. Increased CPA1 may alternately protect against T1D through direct acinar-to-beta cell communication or enzymatic cleavage of proteins in the gut or other tissues. As variants associated with CPA1 levels are also enriched in cell types in the intestine and liver, these tissues may also mediate the role of CPA1 in T1D risk.

Changes in the exocrine pancreas have long been reported in T1D and AAB+ donors, including reduced total pancreas size and volume^42,43^ and decreased trypsinogen, amylase, and lipase^44–47^. Our findings; however, support a causal role for the exocrine pancreas in T1D, which is supported by multiple lines of recent evidence. Genetic association studies have identified risk variants for T1D that affect exocrine-specific gene regulation at numerous loci^1^, which supports a critical role for altered exocrine activity in T1D risk. In addition, a previous study identified a protective role for circulating levels of chymotrypsinogen B1 (CTRB1) in T1D using MR of pQTL data^14^. The gene encoding this protein *CTRB1* maps to a known T1D locus that, based on our results, likely affects acinar cell regulation. As with CPA1, several genes implicated in T1D risk in exocrine cells including *CTRB1/2, CFTR, CEL,* and *GP2* are involved in risk of pancreatitis and/or pancreatic cancer^48–51^, which further supports a potential causal link between inflammation in the exocrine pancreas and risk of T1D.

While the pancreas is the location of autoimmune attack in T1D, our results also support a causal role for other tissues, notably the intestine, in T1D. The digestive tract is heavily exposed to external factors, including likely environmental triggers of T1D^52^. Additionally, the intestine plays a critical role in glucose metabolism by regulating incretin activity through enzymatic cleavage of pro-hormones, which can signal back to the pancreas and modulate beta cell function^53,54^. The gut microbiome has also been implicated in T1D, including potential immune modulation by commensal bacteria^55^. Individual T1D risk loci further highlight a likely role for the intestine in T1D. Most notably, the *FUT2* locus encodes fucosyltransferase 2, an enzyme expressed in intestinal epithelial cells that determines blood group antigen secretor status and is linked to altered risk of viral and bacterial infection^56,57^. This enzyme mediates the transfer of sugar molecules on proteins or lipids^58^. The *FUT2* locus colocalized with amylase and other pancreatic enzyme levels, therefore, it may also affect T1D risk through altered sugar metabolism and glycosylation and/or modulation of enzyme activity in the gut. Digestive enzymes produced in the pancreas may contribute to T1D risk via activity in the gut independently of directly in the pancreas, although the mechanisms are currently unclear. Single cell assays of pancreatic islets have provided insight into processes involved in T1D in the pancreas^34^; however, to date there are no single cell studies looking at intestinal profiles in T1D, which may help clarify the role of the gut in T1D^59^.

One key difference in our study compared to previous efforts is the use of *trans* pQTLs, which can provide additional information beyond *cis* pQTLs to colocalization and MR analyses with the caveat that *trans* signals more often represent indirect effects. To address this, we performed numerous sensitivity analyses to confirm the robustness of instruments including assessing consistency in *cis* pQTLs and both removing outliers and excluding individual variants from instruments. While population-scale proteomics is an invaluable tool for understanding protein regulation in health and disease, studies are often limited to circulating blood due to tissue availability and the large sample sizes required to perform QTL mapping^17,60^. Circulating blood profiles are valuables biomarkers, as they can be collected from any donor, but proteomics analyses in key disease tissues such as pancreas and gut in larger cohorts will be critical for clarifying the mechanistic role of proteins in T1D risk. In addition, two key limitations of current large-scale proteomics platforms, such as the one used in UKB-PPP, are that antibody-based proteomics is unable to detect protein isoforms or posttranslational modifications and only a proportion of the human proteome is assayed^61^. More comprehensive profiling could reveal additional circulating proteins involved in T1D risk.

Preventative treatments for T1D are currently limited, with anti-CD3 monoclonal antibody treatments (e.g. Teplizumab) being one of the only available therapies to delay the onset of T1D^62^. The identification of proteins, cell types, and pathways with evidence for a causal role in T1D, in particularly digestive proteins and systems, provides novel opportunities for therapeutic development outside of immune modulation. Furthermore, the identification of biomarkers of T1D greatly facilitates the identification of likely progressors to T1D for both clinical trials and earlier interventions. This is particularly relevant in the very early stages of disease such as single AAB+ donors, as a minority of these individuals will eventually develop T1D^63^. Overall, our results will enhance efforts to both identify novel therapeutic targets and biomarkers of T1D, as well as studies to understand the molecular mechanisms underlying disease.

## Methods

### T1D GWAS Meta-analysis

We performed a T1D GWAS meta-analysis to use as the outcome in Mendelian randomization analyses, excluding the UKB cohort from the meta-analysis to help ensure independence in sampling between the exposures and the outcome. Briefly, we leveraged summary statistics from our previous T1D GWAS meta-analysis, which included control samples matched for country of origin and genotyping array^1^. Summary statistics from the remaining non-UKB cohorts (**Supplementary Table 1)** were re-meta-analyzed by calculating cohort weights for each variant using the inverse of the variance in each cohort. In total, our T1D GWAS meta-analysis included data from 157,085 individuals (17,497 cases and 139,588 controls). Genetic analyses were approved by the Institutional Review Board (IRB) of the University of California San Diego.

### Colocalization

For each significant pQTL signal, we defined a 2 Mb region flanking the variant with the lowest p-value. Variants in the 2 Mb region for one signal were excluded from being included in regions from other signals for the same protein. Next, we selected all variants in this region included in both the pQTL and T1D GWAS summary statistics. Variant allele frequencies and effect sizes were aligned to the minor allele prior to merging datasets to ensure we are capturing true shared effects between the datasets. Colocalization was performed using the coloc v5.2.3 package with the following default prior probabilities: p1=1×10^-4^, p2=1×10^-4^, and p12=1×10^-5^ ^18^. Since coloc implements Approximate Bayes Factors, no Markov chain Monte Carlo settings were required. For signals with a posterior probability of a shared causal variant (PPH4) greater than 0.8, we additionally calculated the correlation (r^2^) of the lead T1D variant and lead pQTL variant at the shared signal using the 1000 Genomes Project European superpopulation reference panel and PLINK 1.9. If a variant was absent from the reference panel, we used the proxy variant with the strongest LD to the original variant. We considered signals shared if the PPH4 was 0.8 or greater and the r^2^ was 0.5 or greater.

Additionally, colocalization of plasma pQTLs and tissue-specific GTEx eQTLs was performed to assess which tissues regulate protein levels measured in circulating blood. We performed statistical colocalization under the single causal variant assumption, only considering the cis-locus for each protein/gene. We considered signals shared if PPH4 was 0.8 or greater.

### Mendelian Randomization

Instrumental variables were used to assess whether circulating protein levels were causal for T1D. To generate the instrument for each protein, we included the genetic variant with the highest posterior inclusion probability (PIP) from each credible set in fine-mapping data. We created two instruments per protein, one with both *cis* and *trans* QTL signals and one with *cis* pQTL signals only. In each instrument we included only variants with a p-value<1.7×10^−11^ (5×10^-8^/2,922 tested proteins) to ensure strong association with circulating protein levels, which is a key assumption in Mendelian randomization.

Variants from the pQTL instrument and the T1D GWAS meta-analysis described above were harmonized using the TwoSampleMR v0.5.7 package using the provided allele frequencies to determine which alleles are on the forward strand. Only variants found in common between the protein exposure instruments and the outcome GWAS were used in Mendelian randomization analysis. Mendelian randomization was performed using the IVW method within the TwoSampleMR v0.5.7 package to calculate both the direction and the strength of a causal effect between circulating proteins as exposures and T1D as the outcome^17^.

In order to elucidate the contributions of individual cell types on proteins identified as causal from MR, we performed cell type-specific MR using two independent methods. In the first method, we generated instruments from fine-mapped pQTL data as described above but restricted each instrument to variants that overlapped cREs identified in each adult CATlas cell type and subsequently performed inverse variance weighted MR using the TwoSampleMR v0.5.7 package. Individual variants could be found in multiple instruments if cREs were shared across multiple cell types and cell type-specific MR results were considered significantly causal at an FDR < 0.1. In the second method, we used variants from plasma *cis* pQTLs, as described above, and extracted these variants in tissue-specific GTEx expression QTL (eQTL) data to generate ‘cell type-specific’ instruments. For tissues with more than 1 variant, we performed inverse variance weighted Mendelian Randomization and for tissues with less than 1 variant, we performed a Wald ratio using the TwoSampleMR v.0.5.7 package. Genes were considered causal at an FDR < 0.1 after multiple test correcting for the number of unique tissues tested per gene.

### Mendelian Randomization Sensitivity Analyses

We assessed whether MR instruments had a pleiotropic bias by calculating the Egger’s regression intercept using the TwoSampleMR v0.5.7 package, which both provides the regression intercept as well as calculates whether the intercept is significantly different than 0. We considered instruments pleiotropic at an FDR <0.1 and mildly pleiotropic at an uncorrected p-value < 0.05. Next, we used MR-Pleiotropy and REsidual Sum and Outlier (MR-PRESSO) to both assess whether MR instruments contained outliers as well as to re-perform outlier-corrected MR^25^. MR-PRESSO p-values were fit to a Gaussian distribution to be comparable to those computed by the TwoSampleMR package rather than the t-test performed within MR-PRESSO. Finally, we performed leave-one-out analyses using the TwoSampleMR v0.5.7 package, re-assessing MR causal estimates after the removal of individual variants from protein instruments.

### Pathway Enrichment Analyses

Causal proteins were divided into two groups based on their MR effect sizes: protective proteins (MR_beta_ < 0) or risk-inducing proteins (MR_beta_ > 0). Pathway enrichment was performed using Enrichr, which implements a Fisher’s exact test to calculate the probability of a protein’s inclusion in a particular gene set^64,65^. Kyoto Encyclopedia of Genes and Genomes (KEGG) 2021 human gene sets were utilized for pathway enrichment. Multiple test correction was performed on pathways using a Benjamini-Hochberg FDR and pathways were considered significantly enriched at FDR < 0.1.

### Cell Type Enrichment in cREs

We identified annotations enriched for pQTL associations using linkage disequilibrium score regression (LDSC) v1.0.1^26^. First, individual pQTLs were formatted using MungeSumstats with a minor allele frequency threshold of 0.01, chunk size of 500,000, and subset to only include HapMap variants. Next, functional annotations were generated from 110 adult human tissues using single nucleus assay for transposase accessible chromatin sequencing (snATAC-seq) data included in the cis-element atlas (CATlas), using LD data from the 1000 Genomes Project European superpopulation (1KGP EUR)^66^. LDSC was performed on cell type cis regulatory elements (cREs) using a 1 cM window to estimate LD scores from 1KGP EUR for HapMap variants. Finally, partitioned heritability was performed to calculate enrichment of each pQTL in cREs for each cell type^27^. Baseline LD and variant frequencies were calculated from 1KGP EUR reference data. Genomic enrichments and standard errors are reported using the baseline_L0 model. For T1D risk loci that colocalized with causal proteins identified from MR, we used bedtools v2.26.0 to overlap T1D credible set variants with cell type cREs^67^.

### Changes in gene expression in T1D donors

To assess expression levels of pancreatic enzyme genes across diabetes status, we utilized two independent single cell datasets, one generated from single cell RNA sequencing (scRNA-seq) data of pancreatic islets from 65 non-diabetic (ND), autoantibody positive but non-diabetic (Aab+), T1D, or type 2 diabetic (T2D) donors from the Human Pancreas Analysis Program and another generated from single nuclei RNA sequencing (snRNA-seq) of whole pancreas from 32 ND, Aab+ and T1D donors from the Network for Pancreatic Organ Donors with Diabetes (nPOD)^34,68^. For each cell type, we generated normalized gene expression levels for each donor as transcripts per million (TPM), using aggregated raw counts of cells from the donor and GENCODE v38 exonic gene sizes. To assess overall trends in gene expression across all disease groups, we assigned each group a numerical value based on disease progression (ND=1, single Aab+=2, multiple Aab+= 3, recent-onset T1D=4 and long duration T1D=5) and performed DESeq2 using disease progression as a continuous variable including donor sex, scaled age, scaled BMI, tissue source, 10x kit chemistry, and assay (single cell vs. single nuclei) as model covariates. We considered genes significantly up- or down-regulated at an FDR < 0.1. To compare gene expression levels between non-diabetic donors and specific stages of diabetes, we used DESeq2 v1.34.0 to perform differential gene expression analyses in acinar cells in single Aab+, multiple Aab+, recent-onset T1D (T1D < 5 years), or long duration T1D (T1D > 5 years) donors compared to ND controls. We included the covariates specified above and calculated the Wald test statistic with 8 degrees of freedom. Only genes with a minimum of 5 counts in at least 50% of the samples in a condition were tested and included in analyses. We considered genes significantly up- or down-regulated at an FDR < 0.1.

## Data Availability

Summary statistics for the T1D GWAS meta-analysis without UKB samples has been deposited into the NHGRI-EBI GWAS catalogue with accession number GCP000982. Protein QTL summary statistics from the UKB-PPP are available at https://www.synapse.org/Synapse:syn51364943/wiki/622119 and fine-mapping of pQTLs can be found in the following manuscript https://doi.org/10.1038/s41586-023-06592-6. Cell type cREs are available as part of the Cis-Element Atlas (CATlas) http://catlas.org/catlas_hub/.

## Code Availability

All custom code and data analysis pipelines are available at https://github.com/Gaulton-Lab/T1D_protein_biomarkers.

## Acknowledgements

This work was supported by NIH grant DK105554, the Larry L Hillblom foundation, and the Winker Endowment in Type 1 Diabetes Research to K.J.G.

### DCCT/EDIC

The Diabetes Control and Complications Trial (DCCT) and its follow-up the Epidemiology of Diabetes Interventions and Complications (EDIC) study were conducted by the DCCT/EDIC Research Group and supported by National Institute of Health grants and contracts and by the General Clinical Research Center Program, NCRR. The data from the DCCT/EDIC study were supplied by the NIDDK Central Repositories.

### GENIE

The Genetics of Nephropathy, an International Effort (GENIE) study was conducted by the GENIE Investigators and supported by the National Institute of Diabetes and Digestive and Kidney Diseases (NIDDK). The data from the GENIE study reported here were supplied by the GENIE investigators from the Broad Institute of MIT and Harvard, Queens University Belfast and the University of Dublin.

### GoKinD

The Genetics of Kidneys in Diabetes (GoKinD) Study was conducted by the GoKinD Investigators and supported by the Juvenile Diabetes Research Foundation, the CDC, and the Special Statutory Funding Program for Type 1 Diabetes Research administered by the National Institute of Diabetes and Digestive and Kidney Diseases (NIDDK). The data [and samples] from the GoKinD study were supplied by the NIDDK Central Repositories. This manuscript was not prepared in collaboration with Investigators of the GoKinD study and does not necessarily reflect the opinions or views of the GoKinD study, the NIDDK Central Repositories, or the NIDDK.

### T1DGC

This research utilizes resources provided by the Type 1 Diabetes Genetics Consortium (T1DGC), a collaborative clinical study sponsored by the National Institute of Diabetes and Digestive and Kidney Diseases (NIDDK), National Institute of Allergy and Infectious Diseases (NIAID), National Human Genome Research Institute (NHGRI), National Institute of Child Health and Human Development (NICHD), and the Juvenile Diabetes Research Foundation International (JDRF) and supported by U01 DK062418. The UK case series collection was additionally funded by the JDRF and Wellcome Trust and the National Institute for Health Research Cambridge Biomedical Centre, at the Cambridge Institute for Medical Research, UK (CIMR), which is in receipt of a Wellcome Trust Strategic Award (079895). The data from the T1DGC study were supplied by dbGAP. This manuscript was not prepared in collaboration with Investigators of the T1DGC study and does not necessarily reflect the opinions or views of the T1DGC study or the study sponsors.

### T1DGC (ASP/UK GRID)

This research was performed under the auspices of the Type 1 Diabetes Genetics Consortium, a collaborative clinical study sponsored by the National Institute of Diabetes and Digestive and Kidney Diseases (NIDDK), National Institute of Allergy and Infectious Diseases (NIAID), National Human Genome Research Institute (NHGRI), National Institute of Child Health and Human Development (NICHD), and Juvenile Diabetes Research Foundation International (JDRF).

### WTCCC

This study makes use of data generated by the Wellcome Trust Case Control Consortium. A full list of the investigators who contributed to the generation of the data is available from www.wtccc.org.uk. Funding for the project was provided by the Wellcome Trust under award 076113.

### FinnGen

We want to acknowledge the participants and investigators of the FinnGen study.

## Competing Interests

K.J.G has done consulting for Genentech, received honoraria from Pfizer, and holds stock in Neurocrine biosciences. J.C. holds stock in and is employed by Pfizer Inc.

## Author Contributions

R.M.E. contributed to study design, performed data analysis, generated figures, and wrote the manuscript. R.L.M., J.C. and C.W.M. contributed to data analysis. K.J.G. supervised the study, contributed to study design, and wrote the manuscript.

---

**Supplementary Fig. 1:**
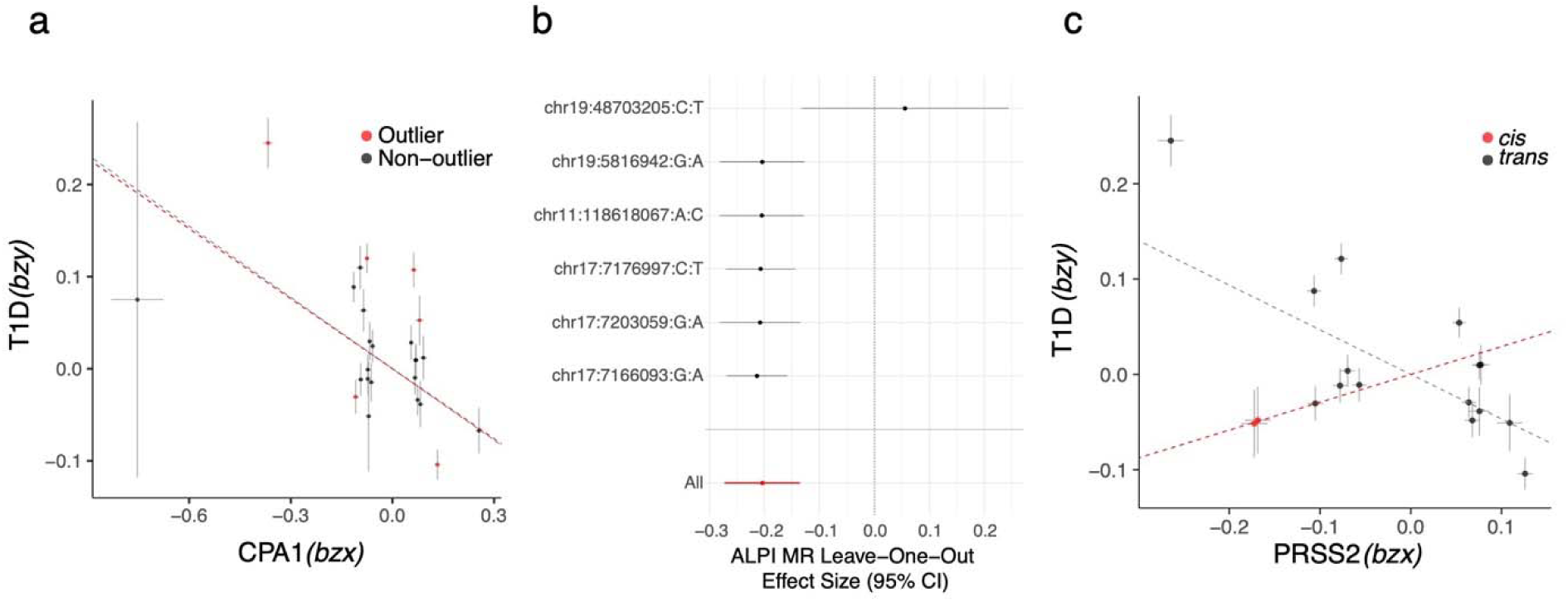
Mendelian randomization sensitivity analyses. **a)** Mendelian randomization effect plots with plasma CPA1 levels as the exposure and T1D as the outcome. Red data points represent outliers detected from MR-PRESSO. Inverse variance weighted Mendelian randomization effect size is represented as a red dashed line for the outlier-removed instrument and as a grey dashed line for the full instrument. Error bars represent standard error of the mean for individual variants. **b)** Leave-one-out Mendelian randomization effect sizes for plasma ALPI levels on T1D. Black data points represent the Mendelian randomization effect size removing each individual variant from the instrument. Error bars represent standard error of the mean of Mendelian randomization effect sizes. **c)** Mendelian randomization effect plots with plasma PRSS2 levels as the exposure and T1D as the outcome. Red data points represent *cis* variants and grey data points represent *trans* variants in the instrument. Inverse variance weighted Mendelian randomization effect size is represented as a red dashed line for the *cis*-only instrument and as a grey dashed line for the *cis* and *trans* instrument. Error bars represent standard error of the mean for individual variants.

**Supplementary Fig. 2:**
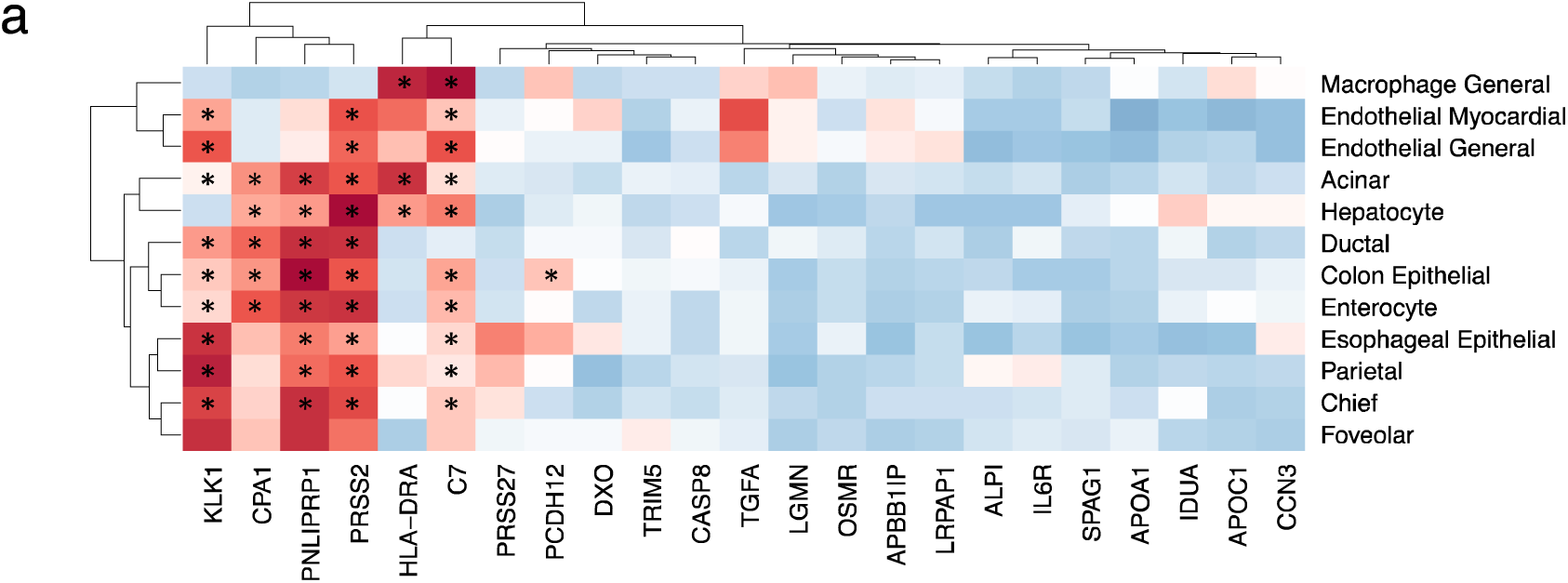
Cell type enrichments of protein QTLs using cell type-specific chromatin accessibility. **a)** Heatmap of LD score regression enrichment -log_10_(p-values) for the top cell types from CATlas. Values are z-scored and scaled per cell type. Asterisks represent significant enrichments (* FDR < 0.1).

**Supplementary Fig. 3:**
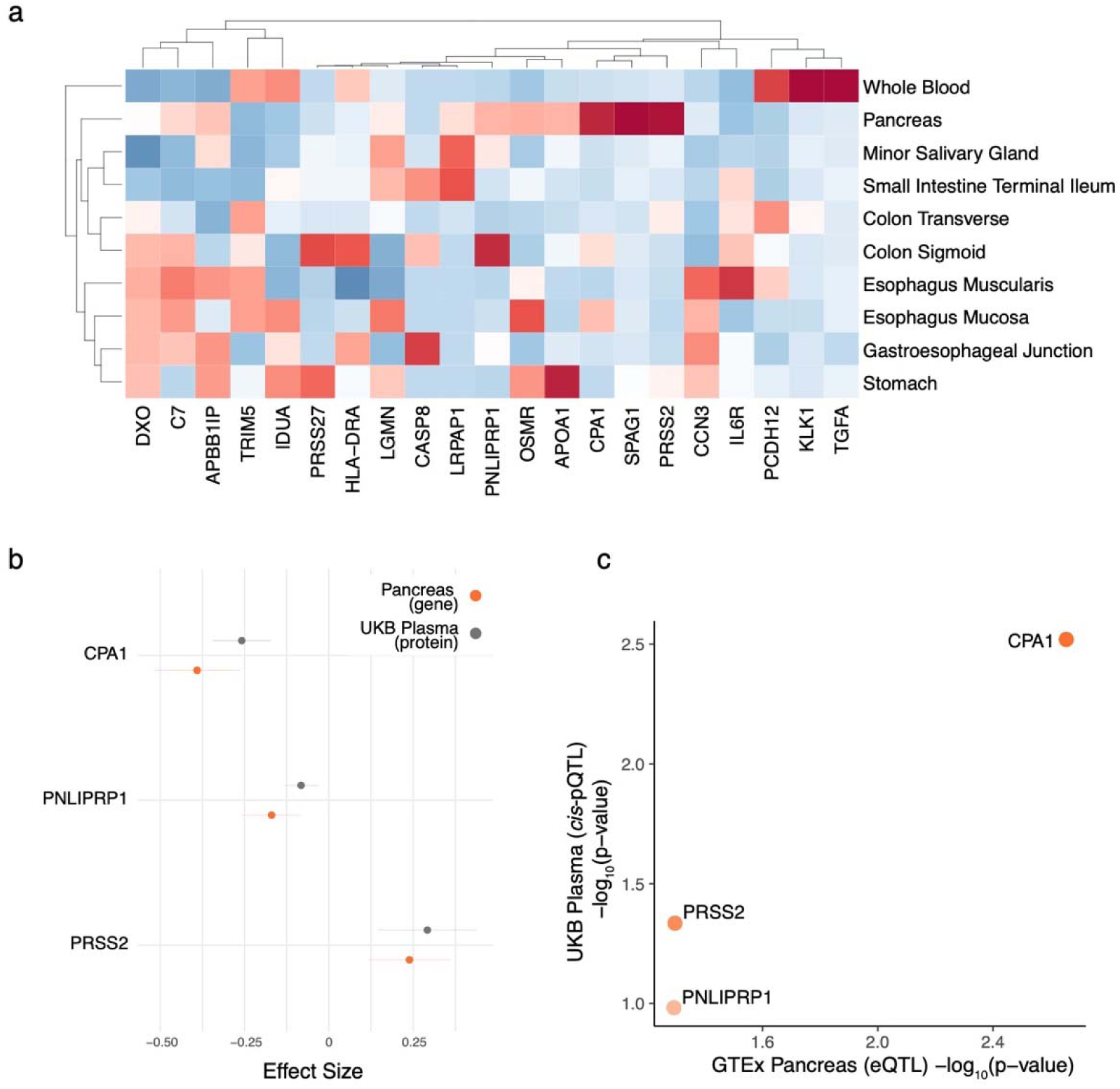
Concordance of GTEx tissue-specific genes and UKB-PPP plasma proteins. **a)** Heatmap showing posterior probabilities of shared causal variants between *cis*-pQTLs and *cis*-eQTLs in select GTEx tissues for 23 causal proteins in T1D. Values are z-scored and scaled per protein. **b)** Mendelian randomization effect sizes of CPA1, PNLIPRP1, and PRSS2 on T1D using *cis*-only plasma pQTLs (grey) or *cis*-only GTEx pancreas eQTLs (orange). Error bars represent standard error of the mean for Mendelian randomization effect sizes. **c)** Mendelian randomization -log_10_(p-values) for CPA1, PNLIPRP1, and PRSS2 on T1D using *cis*-only plasma pQTLs (y-axis) or *cis*-only GTEx pancreas eQTLs (x-axis).

## Notes

### Author Declarations

The Institutional Review Board of the University of California San Diego gave ethical approval for this work

